# Assessment of HeartModel® Automated Left Ventricular Ejection Fraction for Patients with Hypertrophic Cardiomyopathy

**DOI:** 10.1101/2025.10.30.25339136

**Authors:** John Morrissey, Libin Wang, Monica Dehn, Ayan R. Patel, Arsalan Rafiq, Madhavi Kadiyala, Michael Hughes, Ethan Rowin, Martin Maron, Benjamin S. Wessler

**Affiliations:** Department of Medicine, Tufts Medical Center, Boston MA; CardioVascular Center, Tufts Medical Center, Boston MA; Department of Computer Science, Tufts University; Hypertrophic Cardiomyopathy Center at Lahey Hospital and Medical Center, Burlington MA

**Keywords:** Hypertrophic Cardiomyopathy, Transthoracic Echocardiography, Machine Learning, Cardiac myosin inhibitors

## Abstract

**Background:** Cardiac myosin inhibitors (CMIs) have revolutionized care for patients with obstructive hypertrophic cardiomyopathy (HCM), however they are associated with a risk of systolic dysfunction requiring longitudinal echocardiographic monitoring. Machine learning (ML) algorithms applied to echocardiography might expand access to frequent accurate assessment of left ventricular ejection fraction (LVEF). Existing algorithms have not been tested on patients with HCM. We assess the performance of a commercial ML-based LVEF model for patients with HCM.

**Methods:** Single center prospective study of transthoracic echocardiography (TTE) measurements of left ventricular function by Philips HeartModel® (automated) assessment and echocardiographer assessment (standard) for patients with HCM. Assessments of LVEF, end diastolic volume (EDV) and end systolic volume (ESV) were studied across methods and modalities. Median percent differences between measurement methods and correlation coefficients were calculated. Clinical decisions about CMI dosing strategies were modeled around LVEF < 50%.

**Results:** 50 patients with HCM were included. Median age 64 years; 64% were male. Median automated LVEF was lower than standard assessment (55.5% (IQR 9) vs 62.5% (IQR 10), p 0.002, median difference – 8% (IQR 14)). Automated assessment traced larger EDVs and ESVs compared to standard 2D tracings (141 ml (IQR 66) vs 114 ml (IQR 55), p 0.001, median difference + 20% (IQR 35), and 64 ml (IQR 35) vs. 41 ml (IQR 25), p < 0.001, median difference + 43% (IQR 58)). Correlation between automated and standard assessment of LVEF was modest (R^2 = 0.24). Automated assessment identified 11 (22%) patients as having LVEF < 50% vs. 6 (12%) patients identified using standard imaging assessment.

**Conclusions:** For patients with HCM, automated assessments of LV size and function differ significantly from standard assessments, raising concerns about the use of this ML-enabled LVEF software for this patient population and potential application to guiding CMI treatment decisions.

## Introduction

Hypertrophic cardiomyopathy (HCM) is one of the most common genetic heart diseases worldwide, characterized by abnormal increase in left ventricle wall thickness with hypercontractile LV function^1^. Left ventricular outflow tract (LVOT) obstruction is a major therapeutic target^2^ and cardiac myosin ATPase inhibitors (CMI) have recently emerged as a highly effective therapy^3–5^. CMIs reduce myocardial contractility which lower or abolish LVOT gradients resulting in relief of symptoms and enhanced exercise capacity. However, CMI use can be associated with significant reduction in left ventricular ejection fraction (LVEF) which may require dose adjustment or discontinuation of therapy^6^. Consequently, approval of the first CMI, mavacamten, included a Risk Evaluation and Mitigation Strategy (REMS) program to address potential adverse events.

While serial assessments of LVEF are required for determining dose selection for patients on mavacamten, imaging patients with HCM is challenging because of abnormal contours, sometimes massive hypertrophy, and complex Doppler morphologies that are not found during care of patients without HCM. Disease-specific imaging challenges co-exist with more universal concerns related to LV foreshortening, obliquity and poor endocardial border definition.

Automated interpretation of TTEs is increasingly achievable^7–9^ because of advances in computer vision algorithms, processing power, and a massive increase in digital-labeled data. There are now commercial products that enable automated assessment of LVEF from transthoracic echocardiography (TTE). These tools are being advanced to improve accuracy and expand access to high quality image interpretation, which may enable broad reliable safe dosing of mavacamten for patients with HCM.

While ML methods might expand access to accurate TTE interpretation, the performance of automation tools on imaging from patients with HCM has not been tested. Here we assess the performance of a commercially available automated LVEF software model on TTE imaging from patients with HCM.

## Methods

### Study Design

Prospective study of HCM LV size and function assessment. Patients were identified during routine TTE imaging done as part of clinical care. All patients had a clinical diagnosis of HCM and were followed at the Tufts HCM Center (Boston MA). The diagnosis of HCM was made using imaging in patients with a hypertrophied and nondilated left ventricle in the absence of another disease that could produce similar hypertrophy. TTE imaging was performed in standard fashion on Philips EPIQ Ultrasound systems. 3D acquisitions for use with the Philips HeartModel® were performed to obtain automated LV measurements. Left ventricular assessments were made by Philips HeartModel software model (automated), 2) a single board-certified echocardiographer (AR) blinded to the automated assessments (standard). Human expert assessments were done in standard fashion, using overall visual assessment and biplane method of disks (modified Simpson’s rule). Thirteen (26%) studies were interpreted in the context of ultrasound enhancing agents.

### Commercial Automation Software

The Philips HeartModel® automatically detects chamber boundaries, enabling automated measurements of end diastolic volumes (EDV) and end systolic volumes (ESV) and calculation of LVEF. This automation software was trained on thousands of 3D echocardiograms and has shown good performance across a number of diverse datasets^10–14^. To our knowledge, there are no published data regarding its performance on HCM TTE imaging.

### Assessments

The primary comparison in this study is LVEF as assessed by automated software versus standard expert assessment. This assessment was designed to model how CMI dosing decisions are made in clinical practice. Secondary comparisons include comparisons of LV end diastolic volume (EDV) and LV end systolic volume (ESV) obtained by automated methods and standard 2D method using overall visual assessment and biplane method of disks (modified Simpson’s rule). EDV and ESV measurements were extracted from the reports and were compared against values obtained from automated software.

To explore the impact of using automated LVEF assessments to guide CMI dosing decisions we model interpretation-guided treatment decisions around the threshold of LVEF 50%. CMI therapy is discontinued when LVEF drops < 50%. CMI therapy is continued if LVEF >50%.

### Statistics

Continuous variables are expressed as median (interquartile range (IQR)). Categorical data are presented as number (percentage of total study population, (%)). The median percent difference between measurement methods were also calculated. A P value of less than .05 was used to determine statistical significance. Lastly, to assess the relationship between measurement methods, the co-efficient of correlation (R^2) was calculated for each comparison.

## Results

A total of 50 patients with HCM were included in this study. 32 (64%) patients were male and the median age was 64 years (IQR 18.5). Baseline TTE imaging features are shown in **Table 1**. The median interventricular septum thickness was 16 mm (IQR 5.8). Eight (16%) patients had LVOT obstruction at rest. Twenty (40%) patients had LVOT obstruction with Valsalva. Six (12%) patients had LVEF < 50%. Twenty-eight (56%) patients had at least mild mitral regurgitation.

**Table 1.** Baseline features. Demographic and echocardiographic features taken from transthoracic echocardiography reports. LVEF, Left Ventricular Ejection Fraction; IVSd, Interventricular Septum Thickness at end diastole; LVOT, Left Ventricular Outflow Tract. Data are expressed as number positive for a criterion (percentage) or as median (interquartile range, (IQR)). N represents the number of patients.

|  |  |
| --- | --- |
| N | 50 |
| Demographic Features |  |
| Age, median (IQR), years | 64 (18.5) |
| Sex male, n (%) | 32 (64) |
| Race |  |
| African American, n (%) | 8 (16) |
| Asian, n (%) | 2 (4) |
| Caucasian, n (%) | 40 (80) |
| Echo Features |  |
| LVEF, median (IQR), % | 62.5 (10) |
| LVEF ≤ 50%, n (%) | 6 (12) |
| IVSd Thickness, median (IQR), mm | 16 (5.8) |
| Mitral Regurgitation ≥ mild, n (%) | 28 (56) |
| Resting LVOT Gradient, median (IQR), mmHg | 0 (5.3) |
| Valsalva LVOT Gradient, median (IQR), mmHg | 0 (44.3) |
| Resting LVOT Gradient > 35 mmHg, n (%) | 8 (16) |
| Valsalva LVOT Gradient > 35 mmHg, n (%) | 20 (40) |
| Prior Myectomy, n (%) | 4 (8) |
| Apical Aneurysm, n (%) | 3 (6) |

### Automated vs Standard

Median LVEF, EDV, ESV as assessed by each imaging method of analysis are shown in **Table 2**. The median LVEF assessed by automation was lower than standard assessment (55.5% (IQR 9) vs 62.5% (IQR 10), p 0.002, median difference – 8% (IQR 14)). Automated EDV and ESV assessments were larger than manual 2D tracing values (**Table 3**), EDV p 0.001, median difference + 20% (IQR 35) and ESV p < 0.001, median difference + 43% (IQR 58). Correlation between automated and standard interpretation for LVEF, EDV and ESV was modest, with correlation co-efficient, R^2, ranging from 0.24 - 0.41 (**Figure 1**). Eleven (22%) patients were identified as having LVEF < 50% by automation, while six (12%) patients had LVEF < 50% as determined by standard methods. Feature analysis suggests the automation software performance is sub-optimal in the setting of massive hypertrophy, abnormal LV contours and hyperdynamic function **(Figure 3**).

**Table 2.** Median values for each method (Philips HeartModel Software (“Automated”) and standard interpretation (“STD”)). Left Ventricular (“LV”). Data are expressed as median (interquartile range, (IQR)).

|  | **Automated** | **STD** |
| --- | --- | --- |
| LV Ejection Fraction |  |  |
| Median (IQR), % | 55.5 (8.8) | 62.5 (10) |
| LV End Diastolic Volume |  |  |
| Median (IQR), ml | 141 (66.3) | 113.7 (55.4) |
| LV End Systolic Volume |  |  |
| Median (IQR), ml | 64 (34.8) | 41.1 (24.6) |

**Table 3.**
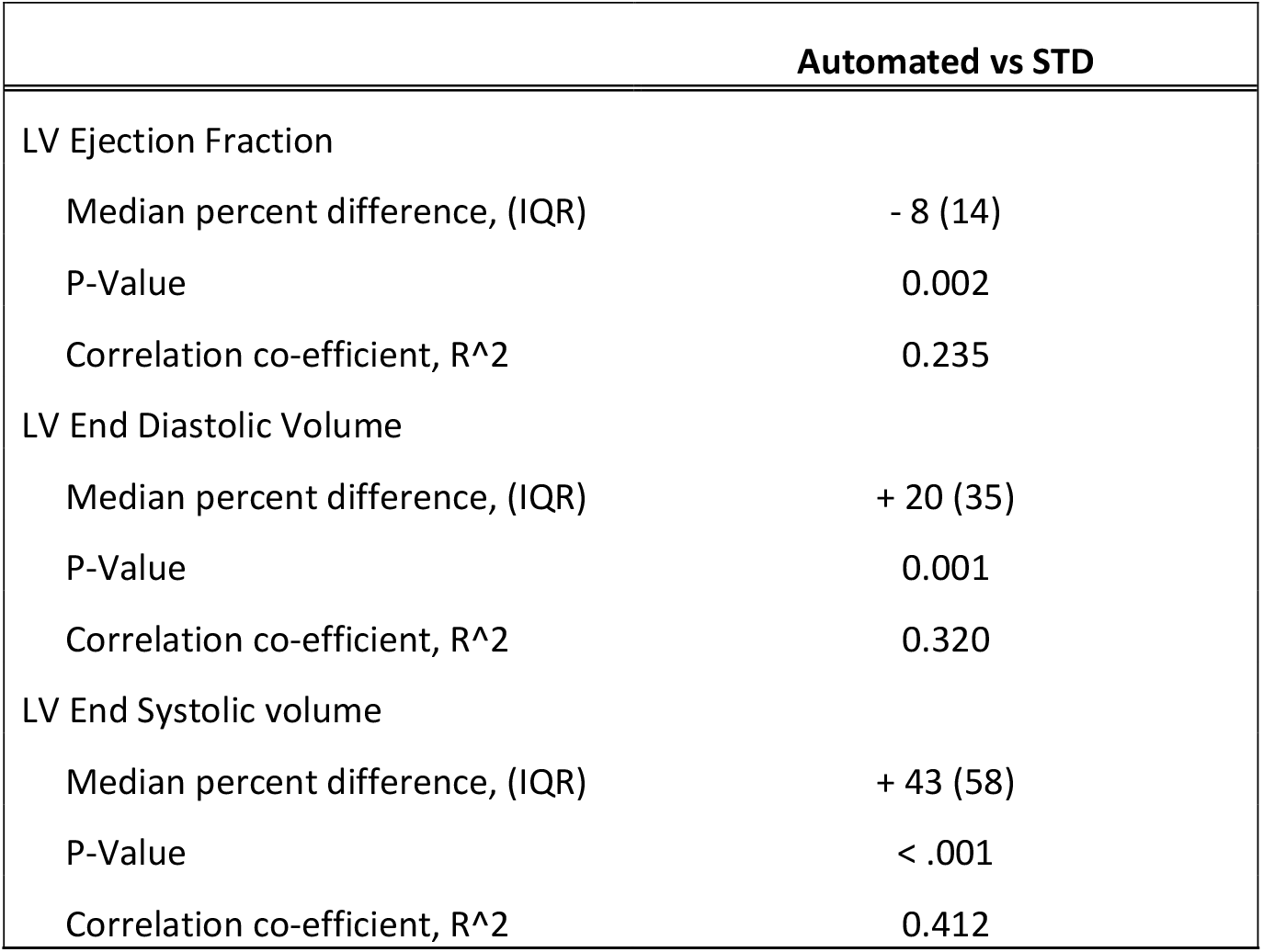
Comparative analysis comparing Philips HeartModel Software (“Automated”) and standard interpretation (“STD). Left Ventricular (“LV”). Data are expressed as median (interquartile range, (IQR)).

**Figure 1.**
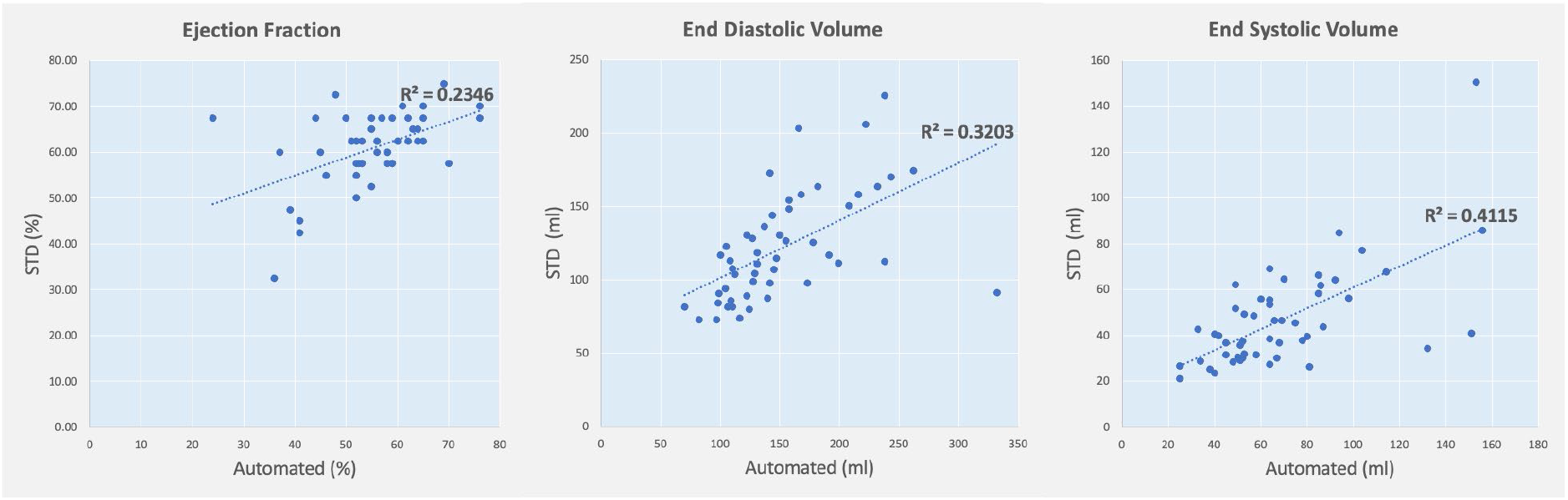
Correlation plots comparing standard methods (“STD”) and Philips HeartModel software (“Automated).Correlation co-efficient (R^2^) displayed for each comparison.

### CMI Clinical Decisions

To model the potential clinical impact of using automated LVEF assessment for patients with HCM, we assessed model performance around an LVEF threshold of 50%. Using blinded echocardiographer (standard) as the ground truth, automated assessment would lead to incorrect dose adjustment or discontinuation for 9 (18%) of these patients. Automated software identified 7 (14%) patients as having an LVEF < 50%, who the standard reported an LVEF > 50%. Additionally, automated software identified 2 (4%) patients as having an LVEF above 50% where the standard reported the LVEF < 50% (**Figure 2, Table 4**). The sensitivity and specificity of the automated software to identify LVEF < 50% was 75% and 86% respectively.

**Table 4.** Table displaying the left ventricular ejection fraction (LVEF) for standard methods (“STD”) and Philips HeartModel Software (“Automated”) and the clinical decision indicated by REMS regarding whether to continue or discontinue mavacamten (“CMI”). LVEF expressed as percentage (%).

| PATIENT | LVEF (%) |  | CMI DECISION |  |
| --- | --- | --- | --- | --- |
|  | *STD* | *Automated* | *STD* | *Automated* |
| 1 | 67.5 | 24 | Continue | Discontinue |
| 2 | 72.5 | 48 | Continue | Discontinue |
| 3 | 67.5 | 44 | Continue | Discontinue |
| 4 | 55 | 46 | Continue | Discontinue |
| 5 | 60 | 37 | Continue | Discontinue |
| 6 | 60 | 45 | Continue | Discontinue |
| 7 | 67.5 | 50 | Continue | Discontinue |
| 8 | 50 | 52 | Discontinue | Continue |
| 9 | 50 | 52 | Discontinue | Continue |

**Figure 2.**
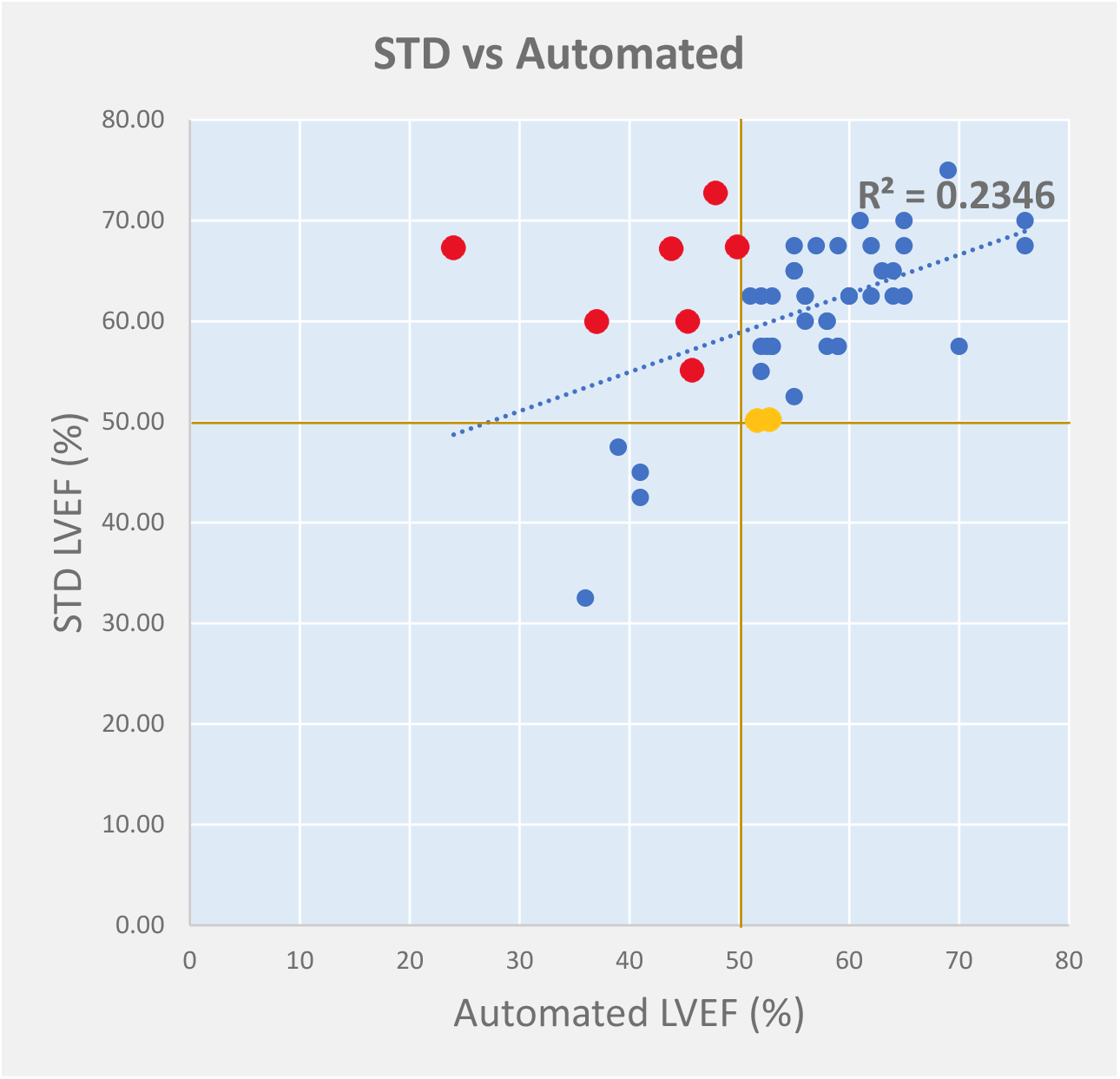
Correlation plot comparing Left Ventricular Ejection Fraction (LVEF) by standard methods (“STD”) with Philips HeartModel Software (“Automated”). Correlation co-efficient (R^2^) displayed. Patients who had an STD LVEF above 50% but an Automated LVEF of 50% or below are represented as red dots. Patients who had a STD LVEF of 50% or below, but an automated LVEF above 50% are represented as orange dots. 50 % LVEF cut off threshold denoted by brown lines.

**Figure 3.**
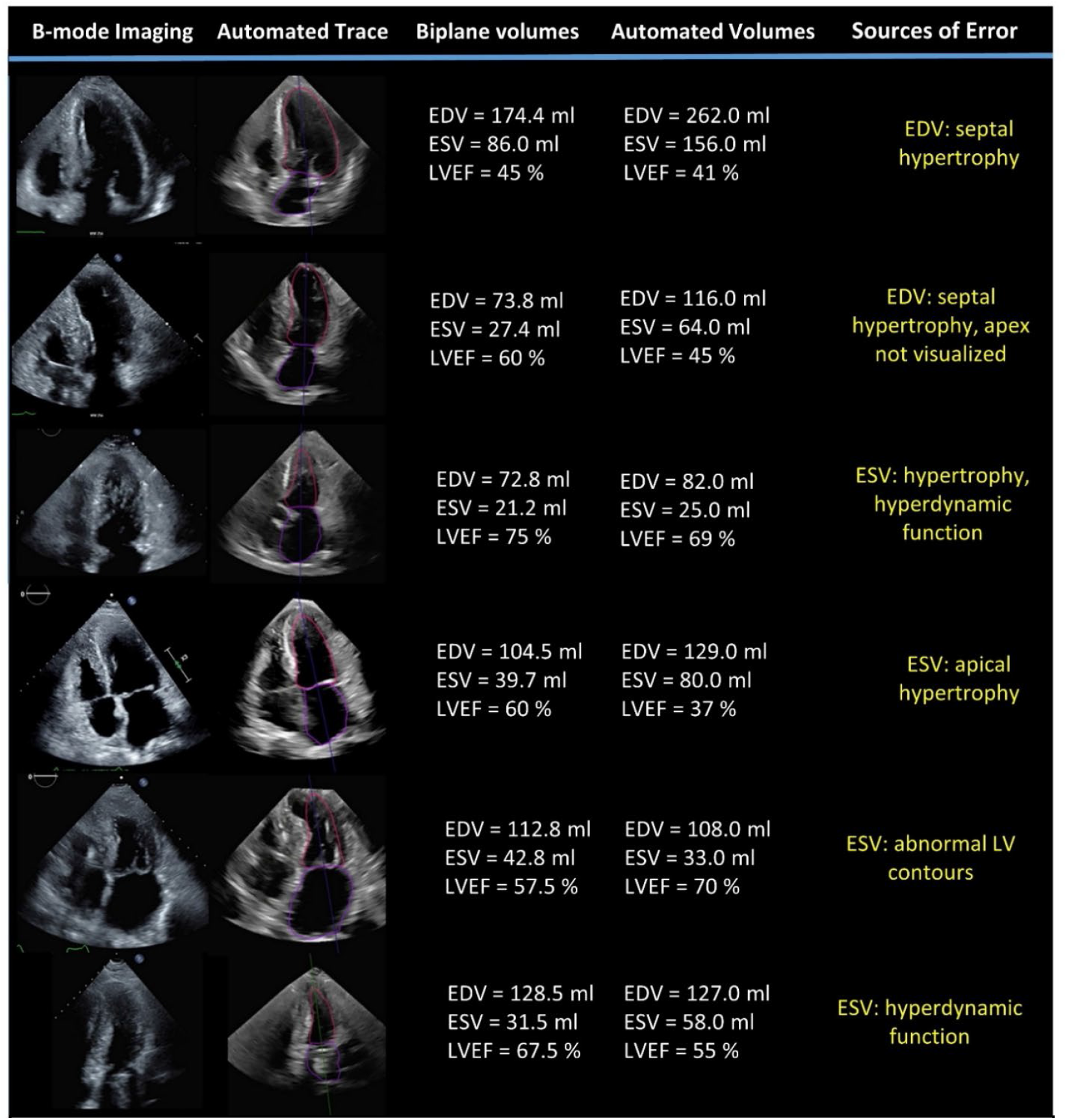
Analysis of automated tracing errors. Representative B-mode imaging frames are shown. Automated tracings represent un-edited outputs from the HeartModel® software. Sources of error were identified by manual review of the automated tracings. EDV, end diastolic volume; ESV, end systolic volume; LVEF, left ventricular ejection fraction.

## Discussion

This study evaluated a commercial automated LVEF tool on HCM imaging with potential to expand access to accurate LVEF assessment. Our main finding was that this automation tool consistently underestimated LVEF and overestimated LV volumes compared to standard expert TTE interpretations. When considering CMI medication dosing around an ejection fraction of 50%, automated assessments would frequently motivate incorrect treatment decisions. These findings raise concerns about this automation tool for HCM image interpretation and suggest that AI-enabled predictions must be rigorously tested before they can be used for this potential use case.

Automated image analysis is an area of substantial interest and has been proposed as a method to expand access to accurate image interpretation. This is predicated on the idea that access to highly trained experts is limited. This concern is particularly salient for patients with HCM, where image interpretation may be challenging outside of dedicated HCM centers with expertise focused on care and management of these patients. While the field of automated image interpretation has experienced substantial recent progress, there are also important concerns about the transportability of models across different datasets and patient populations^15,16^. This study demonstrates that an automation tool that performs well for LV assessments generally^17^ does not appear to perform as well when studied on patients with HCM. This study highlights the need to rigorously test ML models before they are used to inform specific clinical decisions (e.g. CMI dosing).

Reasons for sub-optimal tracing of HCM imaging likely relate to a few issues related to this cardiac phenotype. We identified patterns when reviewing the most error-prone cases: First, HCM hearts often contain abnormal endocardial border contours (e.g. massive hypertrophy) and abnormal patterns of function (hyperdynamic function, hypokinesis from previous myectomy) that were likely under-represented in the original training data for these models. Second, image quality remains a concern for the real-world application of this imaging tool (and many others). While there is growing interest in models that can withhold predictions when there is uncertainty (as good clinicians often do), this field of research remains in its infancy and not yet widely available ^18–20^. While automated border detection can be adjusted by human experts (e.g. sonographers or physicians) and this will likely improve accuracy, this keeps humans in-the-loop and minimizes the potential value of offering opportunity for similar accurate assessments across different levels of expertise.

HCM is a particularly interesting automation use case because of the emerging treatment landscape with CMIs. The mavacamten FDA REMS program requires frequent follow-up TTE assessments of cardiac function (pre-treatment, 4 weeks, 8 weeks, 12 weeks, every 12 weeks) to ensure patients do not develop LV systolic dysfunction as a consequence of mavacamten therapy. Today, these requirements substantially limit medication use outside of specialized centers. These concerns are likely to worsen existing HCM care disparities^21^. In our sample, automated analysis would lead to incorrect CMI decisions in 9 (18%) of patients. Specifically, 7 (14%) patients would have had their CMI discontinued who otherwise would be continued on therapy with standard TTE interpretation and 2 (4%) of patients would have been continued on their CMI when it should have been dose-reduced or discontinued.

There are some limitations that should be considered. First, this test set is of modest size, however it does include patients that represent a full range of LV function. While this study examined a single commercially available model at a single center, it elevates the importance of testing these newer imaging tools in the context of specific use cases prior to deployment.

Third, we used the apparent ground truth from a single echocardiographer at a high-volume HCM center blinded to the automated analysis. While there are well known limitations to the reliability of single readers for LVEF assessment, this study was designed to model how real-world clinical decisions are handled (with a single expert read).

## Conclusion

This analysis does not provide evidence that this automated LVEF software can be used to expand access to accurate HCM image interpretation. Models that are optimized for these patients are needed.

## Data Availability

all data produced in the present study are available upon reasonable request to the authors

## Notes

### Competing Interest Statement

The authors have declared no competing interest.

### Funding Statement

This study did not receive any funding

### Author Declarations

This study was approved by the institutional review board at Tufts Medical Center.

